# Comparative evaluation of clinical and wastewater genomic surveillance for SARS-CoV-2: implications for integrated infectious disease monitoring

**DOI:** 10.64898/2025.12.02.25341504

**Authors:** Tarah Lynch, Andrew Lindsay, Matthew Croxen, Abdul Qureshi, Melodie Barnett, Jennavieve Bu, Theodore Chiu, Ashwin Deo, Paul Dieu, Xiaoli Dong, Christina Ferrato, Stefan Gavriliuc, Fitsum Getachew, Kara Gill, Linnette Imaraj, Roshan Khadka, Petya Koleva, Linda Lee, Vincent Li, Colin Lloyd, Emily McCullough, Stephanie Murphy, Mohammed Abu Naser Mohon, Osahon Obasuyi, Kanti Pabbaraju, Navkiran Randhawa, Silas Rotich, Sandy Shokoples, Sali Sidig, Todd Skitsko, Johanna Skogen, Hilary Trevor, Anita Wong, Christina Yu, Xiaoli Pang, Judy Qiu, Natalie Marshall, Mathew Diggle, Nathan Zelyas, Maria Falsetti, Graham Tipples

## Abstract

**Introduction:** The COVID-19 pandemic demonstrated the need for comprehensive, cost-effective surveillance systems integrating multiple data streams. This study directly compares SARS-CoV-2 genomic data from wastewater-based surveillance (WBS) and clinical diagnostic testing (CDT) to evaluate lineage diversity, detection timing, and persistence patterns that could inform integrated surveillance strategies.

**Methods:** We analyzed SARS-CoV-2 genomic data from Alberta, Canada (July 2022 – March 2025), encompassing 13 municipal wastewater treatment plants covering 80% of the provincial population and clinical samples from provincial diagnostic testing. Clinical samples (n=28,610) and wastewater samples (n=1,685) were sequenced using a tiled amplicon approach. We compared lineage richness over time, lead time for first detection using collection dates and explored four additional wastewater metrics: abundance at first detection, peak abundance, time to peak abundance, and total time detected. The comparison grouped lineages into those found only in WBS and those that were seen in both WBS and CDT.

**Results:** Of the 2,586 unique lineages identified over the study period, 1,588 (61.1%) appeared exclusively in WBS, 42 (1.6%) only in CDT, and 956 (36.9%) in both systems. WBS consistently demonstrated higher monthly lineage richness (95-660 lineages) compared to CDT (23-160 lineages). While WBS detected lineages an average of almost 12 days earlier than CDT, the most frequent pattern showed CDT detection first by 8 days, indicating substantial variability. Lineages detected in both systems showed significantly higher initial relative abundance, peak relative abundance, longer persistence, and delayed time to peak compared to WBS-only lineages (all p<0.0001).

**Conclusions:** WBS and CDT provide complementary surveillance capabilities with distinct strengths. Rather than relying on “*first detection”* as an early warning metric, integrated surveillance should prioritize concordance patterns and abundance metrics that indicate lineages with sustainable transmission potential. These findings support developing surveillance frameworks that strategically combine population-level WBS monitoring with case-linked CDT data for more effective public health response.

**KEY MESSAGES:** *What is already known on this topic:*

- WBS can detect SARS-CoV-2 and other pathogens at the population level and has been described as an early warning system, while CDT provides case-linked genomic data but may be biased towards symptomatic, high-risk or healthcare-seeking individuals.
- Both surveillance methods were used during the COVID-19 pandemic, but it remains uncertain how to best integrate WBS and CDT genomics and which WBS metrics are most informative.

*What this study adds:*

- This study provides the first direct comparison of SARS-CoV-2 genomic data from WBS and CDT processed by a single laboratory over nearly three years.
- WBS consistently captured greater lineage diversity than CDT, but “first detection” showed substantial variability, with lineages frequently detected in clinical samples before wastewater.
- Lineages identified in both surveillance systems showed distinct signatures: higher peak abundance and longer persistence, suggesting these concordance patterns are more reliable indicators than timing alone.

*How this study might affect research, practice or policy:*

- Integrated surveillance frameworks should prioritize monitoring concordance between WBS and CDT, using abundance and persistence metrics to identify lineages with significant transmission potential.
- The complementary strengths of these systems support their combined use for cost-effective surveillance with WBS monitoring population trends and CDT providing clinical context for targeted interventions.

## INTRODUCTION

The COVID-19 pandemic caused by the severe acute respiratory syndrome coronavirus 2 (SARS-CoV-2) accelerated our need for surveillance systems that are cost-efficient, resilient, and adaptable. Clinical diagnostic testing (CDT) is essential for patient management, monitoring outcomes, isolating pathogens (cultured or genomically) for advancing characterization, transmission analysis, and tracking pathogen evolution (1–3). Wastewater-based surveillance (WBS) offers a stable population-level signal even when clinical testing wanes. Combining complementary data streams of CDT and WBS may be the way forward to balance unpredictable or limited resources for public health priorities (4), however the direct comparison of the genomics role has not been fully understood.

This study addresses this gap by directly comparing genomic data from CDT and WBS to evaluate diversity, persistence, and early detection signals. The algorithms for molecular testing evolved through the pandemic and into the post-pandemic era. Quantitative PCR (qPCR) assays were developed for fast detection and relative quantification of SARS-CoV-2 viral load (5). SARS-CoV-2 was subsequently integrated into laboratory-developed and commercial respiratory multi-pathogen panels for CDT while the single target qPCR assay continued to be used for quantifying community trends in WBS (6). Whole genome sequencing quickly gained value to characterize mutations across the full virus genome for lineage designation. In CDT genomics produced a consensus sequence to be shared nationally and internationally to track viral evolution and inform lineage designation (3,7). In WBS, a similar full genome approach was taken to estimate SARS-CoV-2 lineages and their relative abundance, however consensus sequences were not produced (8).

Genomics has proven a valuable tool in outbreak investigations and pathogen evolution studies. During the COVID-19 pandemic, global sequencing of clinical samples enabled variant designation, pathogenicity studies, and vaccine design through data sharing in public repositories. Evidence suggests that WBS can predict clinical burden increases 5-63 days before clinical indicators emerge and detect variants of concern up to 14 days earlier than CDT (8–11). Yet integration of WBS and CDT for effective public health response remains uncertain. The WBS metrics most informative for public health decision-making or understanding disease dynamics need to be identified.

The “first detection” has often been treated as evidence for use of WBS as an early warning system, but it is not obvious when this metric could signal actionable risk, as this statement is often retrospectively used. Additional variables affecting the use of CDT and/or WBS include financial considerations, genomics capacity, and the specific goals of the surveillance program.

This study had three primary objectives: (1) to compare lineage diversity patterns between WBS and CDT, (2) to evaluate the reliability of “first detection” as an early warning metric for WBS, and (3) to identify WBS metrics that correlate with lineages detected in CDT, potentially indicating greater transmission or clinical significance.

## MATERIALS AND METHODS

### Sample population

This study included CDT and WBS across the province of Alberta, Canada from July 1, 2022 – March 31, 2025, when the Provincial Public Health Laboratory (ProvLab) was conducting both for SARS-CoV-2. This timeframe covers the height of the Omicron wave, through the closure of community assessment centers (March 31, 2023) and into the new paradigm of rapid antigen kits for self-testing at home and more relaxed quarantine guidelines. During the pandemic, molecular testing for SARS-CoV-2 was added to commercial respiratory pathogen panels, thus testing for symptomatic and high-risk individuals continued. A limited amount of community testing for SARS-CoV-2 also occurred through the Sentinel Practitioner Surveillance Network (SPSN) (12).

Due to fluctuations in the number of SARS-CoV-2 positive CDT samples throughout the study period, it was not feasible to sequence all samples. Selections were made to prioritize patients admitted to care facilities, outbreaks, and a selection of geographically and age diverse community-based patients across Alberta. This selection skewed the sequenced CDT samples to include more severe infections.

### CDT Protocol

SARS-CoV-2 samples were collected by flocked nasopharyngeal swab administered by a health professional and collected in universal or viral transport media. Multiple testing platforms were used for the initial diagnosis (qPCR assay) and resulted to the patient/healthcare provider; samples with Ct<30 was reflexed to the Genomics section at ProvLab for whole genome sequencing using the Freed primers (v5) (13). Illumina libraries were prepared using the DNA Prep Kit (Illumina, San Diego, USA) while the Nanopore libraries were prepared with the Ligation Sequencing Kit LSK110 (Oxford Nanopore Technologies, Oxford, UK). Whole genome sequencing was performed once or twice per week on either the MiniSeq Mid Output 300 Cycle Kit (Illumina) or the MinION R9.4.1 flow cell (Oxford Nanopore Technologies). Frequency of sample processing was dependent on positive qPCR sample volumes.

Genomic data was analyzed using the ARTIC protocol [https://artic.network/viruses/sars-cov-2/sars-cov-2-bioinformatics-sop.html], with automation scripts by the Connor Lab [https://github.com/connor-lab/ncov2019-artic-nf]. Additional quality assurance metrics were produced using ncov-tools [https://github.com/jts/ncov-tools]. All clinical samples were re-analyzed for this study using Pangolin (v 4.3.1), Pangolin-data (v.1.34), and UShER (v 0.6.3) for lineage assignment. Consensus sequences were submitted to the National Microbiology Laboratory Branch for national surveillance, and are available in GISAID, and the Canadian VirusSeq repositories.

### WBS Protocol

Thirteen samples were collected from 13 wastewater treatment plants located in 11 municipalities across Alberta, covering approximately 80% of the population. The municipalities included were: Grand Prairie, Fort McMurray, Jasper, Fort Saskatchewan, Edmonton, Red Deer, Banff, Calgary (3 facilities), High River, Medicine Hat, and Lethbridge. Samples were sequenced weekly, full genome amplification of SARS-CoV-2 was done using the ARTIC primer scheme (v 5.3) [https://github.com/quick-lab/SARS-CoV-2] primers and sequenced on the Illumina MiniSeq platform. Sequencing data were analysed with the same artic and ncov-tools pipelines, however instead of consensus sequence generation, BAM files were analyzed with Freyja [https://github.com/andersen-lab/Freyja] using default settings with a minimum relative abundance of 0.1%. All samples were re-analyzed for this study using the latest versions, Freyja (v1.5.1) and UShER (v 0.6.3).

### Statistical analysis

Analyses were completed using R (v4.3.0) with the tidyverse package suite (14). Lineage richness was defined as the total number of unique Pangolin lineages per month. First detection lead time was calculated as the difference in collection dates between first appearance in WBS and CDT for each lineage, with negative values indicating earlier clinical detection.

Given the non-normal distributions of abundance and persistence datea, comparisons between lineages detected exclusively in WBS versus those found in both systems used the Wilcoxon rank-sum tests. A p-value threshold of 0.01 was used for statistical significance.

### Ethics

This study was reviewed and approved by the Conjoint Health Research Ethics Board at the University of Calgary (Ethics ID: REB21-0877_REN4). All consensus sequences from CDT are available through GISAID and the Canadian VirusSeq portal.

### Funding

This work was supported by the Public Health Agency of Canada Integral Genomic Innovations, grant number 2223-HQ-000215 and the Public Health Agency of Canada Wastewater Surveillance Fund, grant number 2324-HQ-000049.

## RESULTS

### CDT testing volumes

Alberta maintained a high proportion of genomic sequencing of SARS-CoV-2 through the pandemic and beyond (Figure 1). The criteria for successful genomic sequencing were based on an empirical threshold qPCR Ct <30 and 85% coverage of the full genome length. The positivity rate of CDT qPCR tests was between 1 – 25% with peaks during high testing periods and a gradual decrease over the study period. A decrease in testing volumes can be seen from late 2022 to early 2023 when assessment centers closed and at-home rapid testing became available (Figure 1A). The testing volumes then begin to mirror seasonal respiratory patterns as SARS-CoV-2 was added to multi-target respiratory pathogen panels. During the height of the Omicron wave (mid-late 2022), the proportion of genomics were at their lowest (30-40%) as test volumes exceeded capacity. The proportion of successful genomics testing increased to 80% but then plateaus between 50-60% as both qPCR and genomics volumes decrease into the post-pandemic era (Figure 1B).

**Figure 1.**
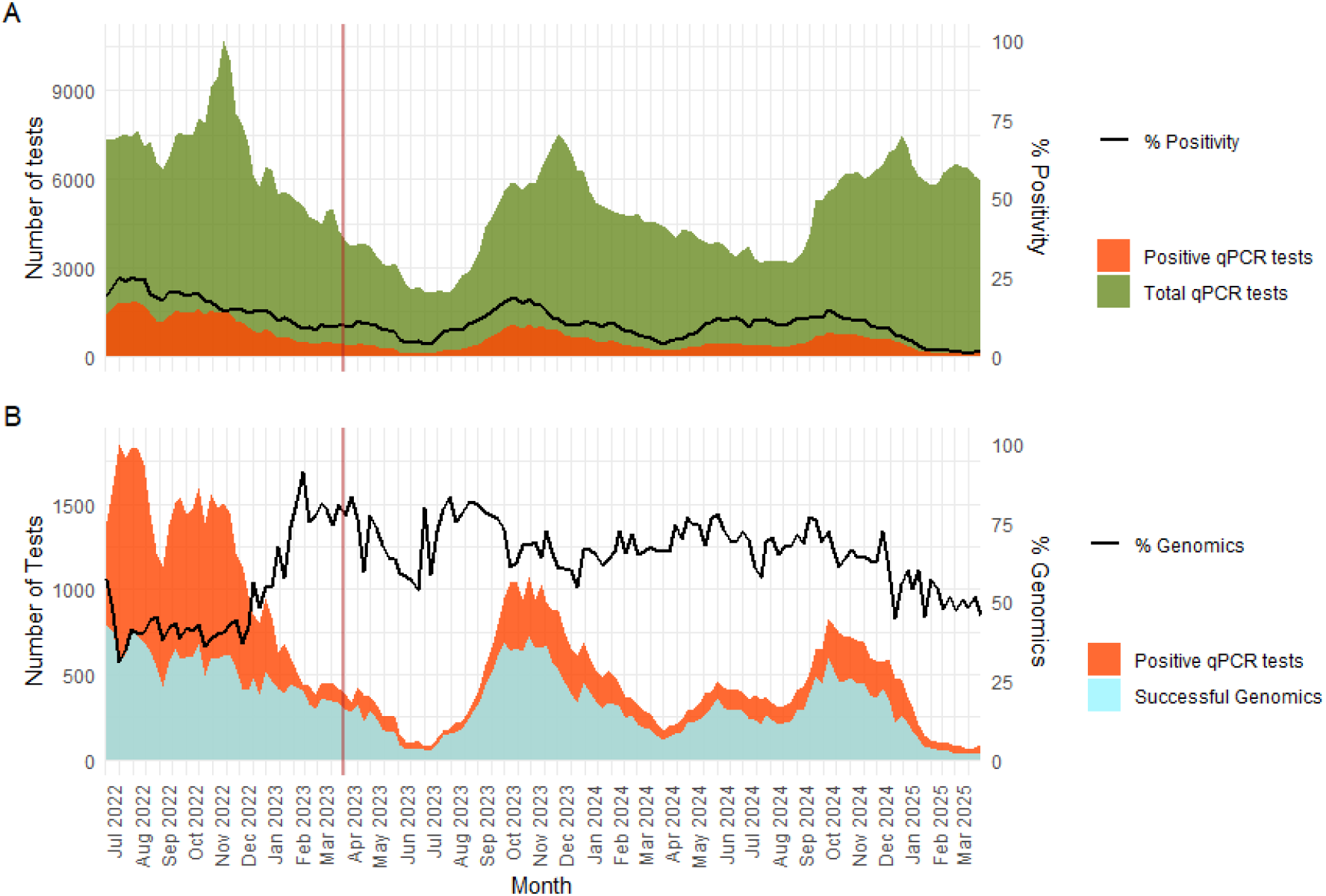
Clinical qPCR and genomic testing volumes and positivity rates. Panel A compares the total number of SARS-CoV-2 qPCR tests (green) to the subset with positive results (orange) and % positivity (black line). Panel B is the proportion of successful genomic sequencing (blue) as a proportion of positive qPCR tests (orange) and the % of samples successfully sequenced (black line). The red vertical line denotes closing of community assessment centers on March 31, 2023.

### Lineage diversity patterns across surveillance systems

Over the study period there was a total of 1588 (61.1%) unique lineages found in WBS only, 42 (1.6%) lineages found only in CDT, and 956 (36.9%) identified in both datasets. Monthly lineage richness was calculated for both WBS and CDT to assess temporal variation in diversity (Figure 2). CDT genomic test volumes ranged from 63 – 2,947 per month, compared to 28 – 52 WBS samples. WBS exhibited substantially higher lineage richness throughout the study period (95 – 660 lineages/month) compared with CDT (23 – 160 lineages/month). WBS richness peaked in early 2023, declined through late 2023, and rose again in early 2025, whereas clinical richness remained lower and showed only modest fluctuations. Overall, WBS consistently recovered greater lineage diversity than CDT.

**Figure 2.**
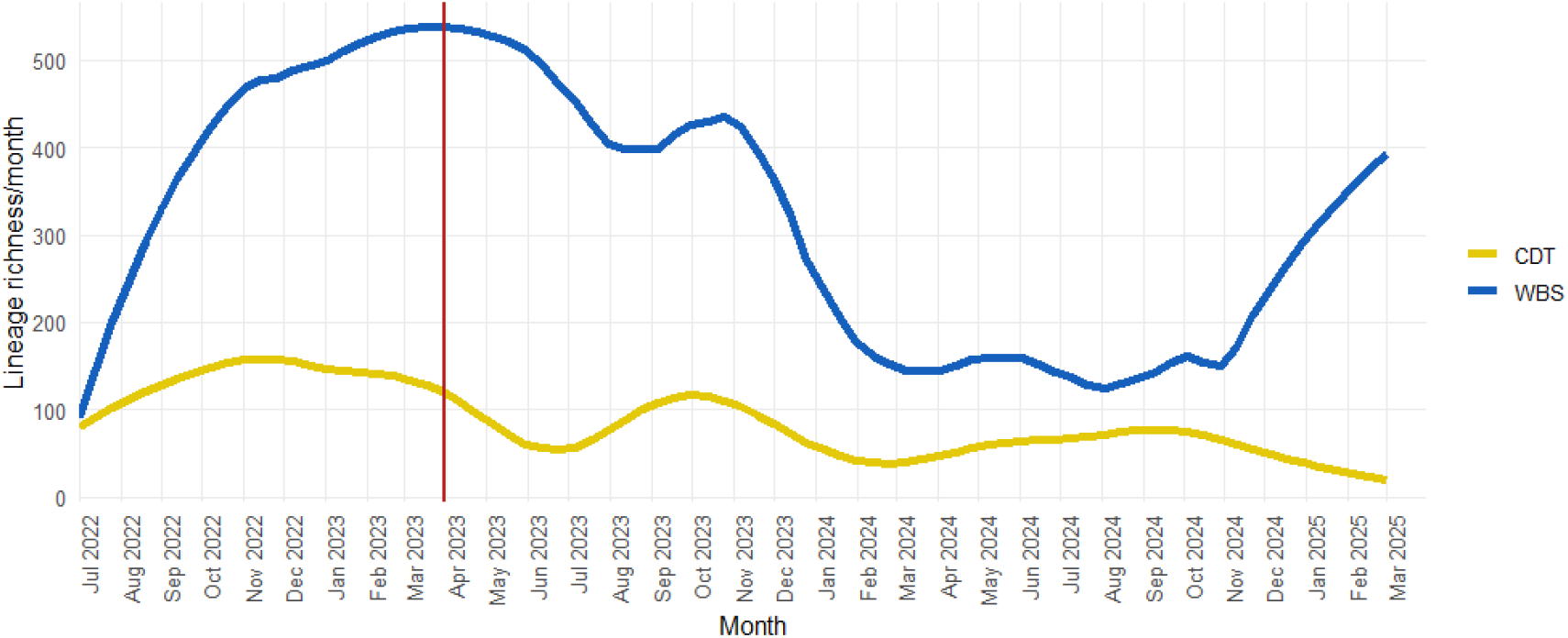
Lineage richness over time for WBS (blue) and CDT (yellow). The red vertical line denotes the closing of the community assessment centers. Lineage richness is calculated as total unique lineages per month.

### Lineage first detection comparison

We calculated the lead time between first detection in WBS and CDT using collection dates for the 956 lineages detected in both datasets (Figure 3). Collection dates were used because although processing times for WBS and CDT are similar (7-14 days), the differing reporting pathways make reported dates unsuitable for comparison. Lead times ranged from −612 to 767 days, with negative values indicating earlier CDT detection (n=385) and positive values indicating earlier WBS detection (n=559); 11 lineages were detected on the same day. The distribution had a mean lead time of 11.9 days (WBS earlier) but a mode of −8 days (CDT earlier), reflecting substantial variability in which system detected a lineage first.

**Figure 3.**
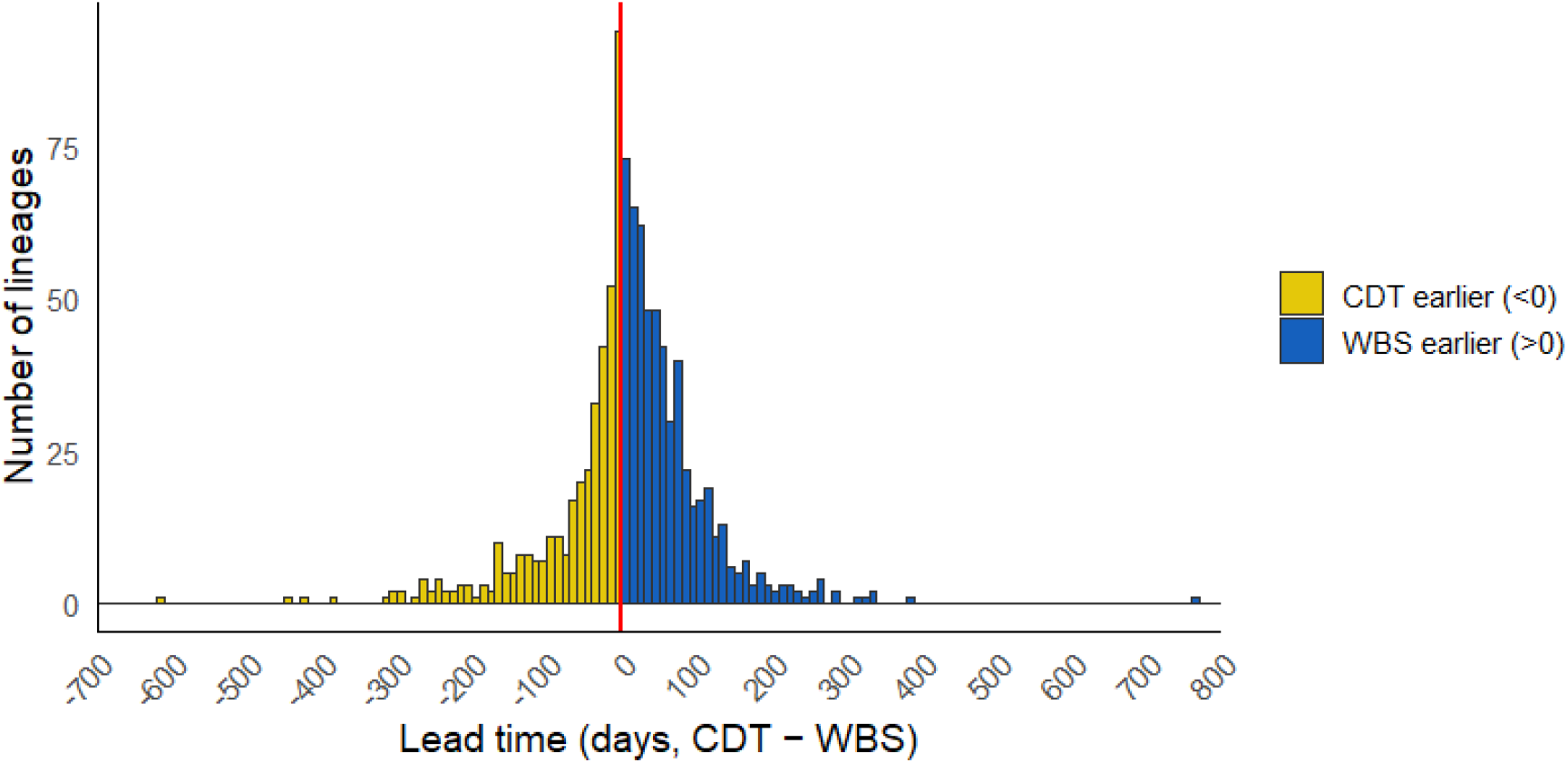
Frequency and timing of first lineage detection in WBS or CDT. Of lineages present in both datasets, lead time is the difference in days between first detection in WBS and CDT using the collection dates. Lead time is displayed on the x-axis: < 0 days are lineages detected in CDT first, 0 = lineages first detected same day, and >0 days are lineages first detected in WBS.

### WBS metrics to identify patterns of lineages found in WBS or both

To assess whether WBS metrics correlated with clinical detectability, we compared lineages observed exclusively in WBS with those found in both WBS and CDT (Figure 4). Among the 1,588 wastewater-only and 956 shared lineages, all four metrics: abundance at first detection (Fig. 4A), maximum abundance (Fig. 4B), time to peak abundance (Fig. 4C), and total time detected (Fig. 4D) – all differed significantly (Wilcoxon, p < 0.0001). Lineages detected in both WBS and CDT appeared at higher initial abundance, reached higher abundance peak, took longer to reach maximum abundance, and persisted longer in wastewater. In contrast, wastewater-only lineages were generally lower in abundance, peaked sooner, and were detected for shorter durations.

**Figure 4.**
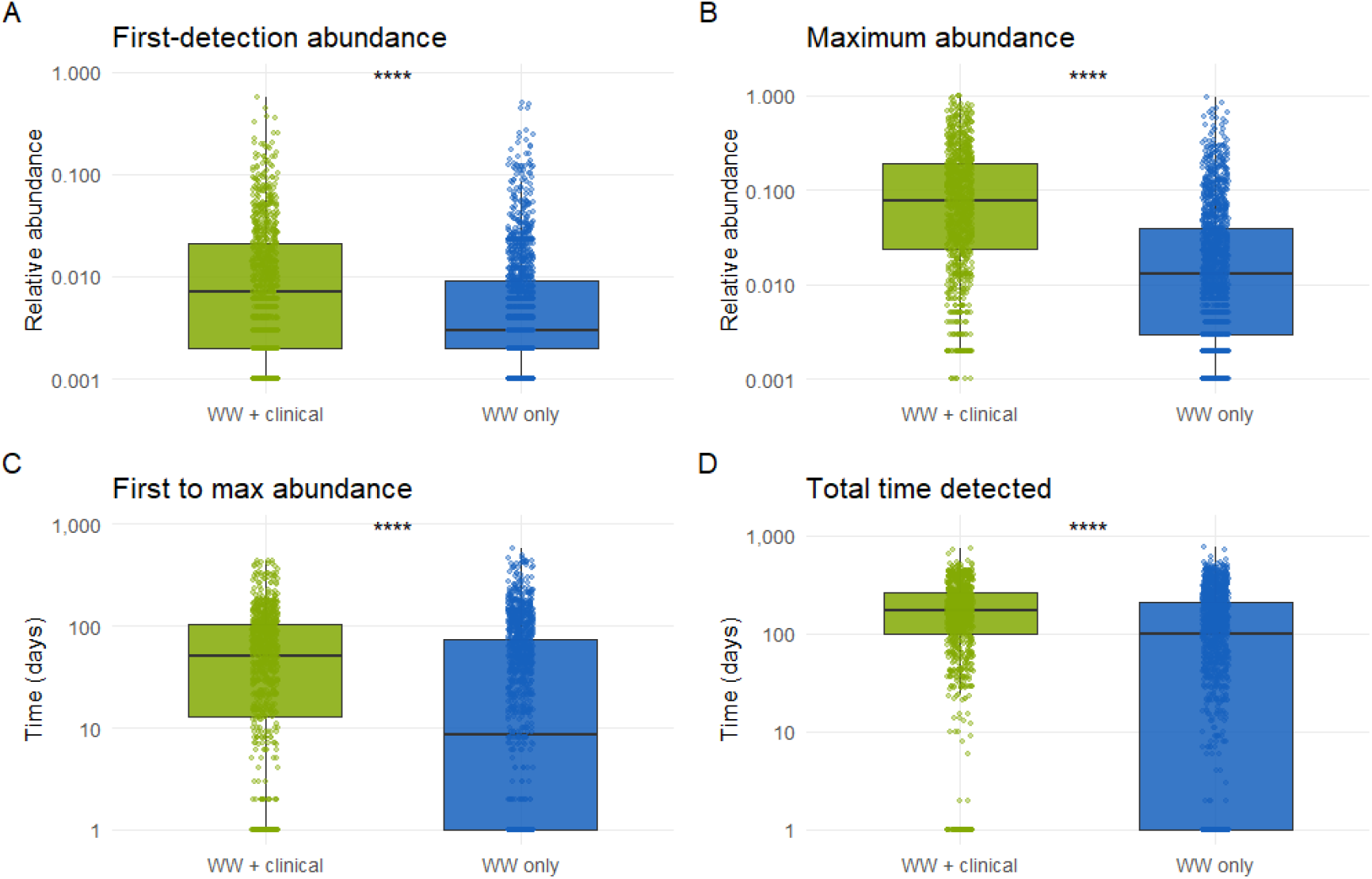
Exploring WBS data to compare lineages exclusively in WBS (WW only, blue) against lineages found in both WBS and CDT (WW + Clinical, green). Total number of lineages in WW only = 1588, WW + Clinical = 967. All comparisons used Wilcoxon rank sum test (**** = p < 0.0001). (A)= relative abundance at first detection, (B)= maximum abundance reached. (C) = length of time from first detection to maximum abundance, (D) = total time detected in WBS. All y-axes are displayed on the log 10 scale.

## DISCUSSION

This study provides the first detailed comparison of SARS-CoV-2 genomic data from WBS and CDT processed through a single laboratory over an extended timeframe spanning multiple pandemic phases. Our findings demonstrate that these surveillance approaches capture different dimensions of viral circulation and should be integrated strategically rather than viewed as redundant or competing systems.

WBS and CDT methods have unique benefits and limitations for infectious disease surveillance (summarized in Table 1). WBS is non-invasive, provides community-level coverage, and includes all severity levels of infection regardless of healthcare access. The purpose of CDT is individual-level results to track symptoms and outcomes required for patient management. While both approaches proved valuable during the pandemic, optimal integration strategies for public health action remain unclear.

Our analysis demonstrates that WBS consistently captured greater lineage diversity than CDT throughout the study period. Despite CDT’s bias towards higher-risk groups and more severe infections, the significant overlap between systems (36.9% of lineages detected in both) suggests CDT remains representative of predominant circulating virus. This finding gains support from our observation that lineages detected in both systems showed higher relative abundance and greater persistence in wastewater, potentially reflecting increased viral shedding associated with symptomatic or severe illness, though viral shedding kinetics are multifactorial (15).

**Table 1.** >Overview of CDT and WBS genomics for infectious disease surveillance.

| Variable | Wastewater-Based Surveillance (WBS) | Clinical Diagnostic Testing (CDT) |
| --- | --- | --- |
| Scope | Population-level coverage, capturing asymptomatic to severe infections. | May be biased to symptomatic, high-risk, and healthcare-seeking individuals. |
| Early Detection | Can detect variants earlier but clinical relevance may be unknown. | Detection may lag due to restricted testing but infections with high morbidity/mortality identified early (i.e. hospital cases). |
| Cost | Laboratory cost equivalent per test, monitors larger urban populations. | Laboratory cost equivalent per test, covers fewer individuals. |
| Lineage Data | Shows broader diversity and longer detection windows. | Provides high-resolution data linked to clinical outcomes. |
| Pathogen Isolation | Complex polymicrobial samples – culture isolation possible but increased labour cost. Difficult to produce consensus sequences for data sharing (currently not done, not impossible). | Pathogen culture isolation possible. Genome data shared to public repositories for global phylogenetic placement and further research and characterization. |
| Limitations | No direct link to severity or outcomes, sewer infrastructure and protocol variability. Data may be missed by diaper users. | Sampling bias, lower population coverage, and limited asymptomatic detection. |

A major finding challenges the widely held assumption that WBS serves as an early warning system for emerging lineages. While WBS detected more lineages earlier on average (mean = 11.9 days), the most frequent pattern showed CDT detection occurring first (mode = −8 days) with the distribution stretching much longer. This substantial variability suggests that “first detection” alone is not a reliable metric for WBS-based early warning systems. Instead, concordance between WBS and CDT appears more informative, as share lineages demonstrated higher abundance and persistence, characteristics that may indicate greater transmission potential and clinical significance.

These findings support developing surveillance strategies that leverage the complementary strengths of both approaches. WBS excels at monitoring population-level trends and can inform community-wide interventions such as public health messaging, vaccination campaigns, and preventative measures (16,17). CDT provides case-linked data crucial for clinical interventions and detailed pathogen characterization. Integrating these approaches enables public health programs to balance broad population monitoring with targeted clinical responses in a cost-effective manner.

Several limitations warrant consideration. Different primer schemes between WBS (ARTIC v5.3) and CDT (Freed v5) may have introduced bias affecting genome coverage and lineage designation, though all met established quality standards. The study did not examine clinical factors such as vaccination status or symptom severity, which may influence lineage diversity and circulation patterns. Third, collection dates across WBS sites were often recorded as a single day each week and therefore may impact the lead time calculations by +/-up to 3 days compared to CDT samples where collection dates are recorded to the minute.

Future studies should integrate more comprehensive clinical variables with WBS data to develop predictive models that better utilize both surveillance streams. Research examining the relationship between WBS data and clinical outcomes could enhance our understanding of which WBS parameters best predict significant transmission events. Modelling could also determine optimal sampling strategies and testing requirements for CDT based on population and pathogen transmission dynamics. Long-term studies across different pathogen systems would help validate these findings beyond SARS-CoV-2.

## CONCLUSIONS

This study demonstrates that WBS and CDT provide complementary rather than redundant surveillance capabilities for SARS-CoV-2. Rather than relying on “first detection” as an early warning metric, integrated surveillance frameworks should prioritize concordance patterns and abundance metrics that identify lineages with sustainable transmission potential. WBS consistently captures greater lineage diversity and provides stable population-level monitoring, while CDT offers essential clinical context for targeted interventions.

## Data Availability

All clinical data in the present study are available through GISAID and the Canadian VirusSeq portal

## ACKNOWLEDGEMENTS

We would like to acknowledge additional contributions from Qiang Jiang, Melissa Wilson, Kimberly Tang, Poonam Patel, Seema Kailey, and Sydney Rudko for their collaborations.

